# A participatory action research study to explore adolescents’ experiences of adverse childhood experiences (ACEs) through creative workshops: a protocol

**DOI:** 10.1101/2023.12.21.23300349

**Authors:** Isabelle Butcher, Anna Mankee Williams, Siobhan Hugh-Jones, Paul Cooke, Ben Teasdale, Nicola Shaughnessy, Gabriela Pavarini, Lindsay Smith, Kamaldeep Bhui

## Abstract

**Introduction:** Adverse Childhood Experiences (ACEs) are associated with poor mental health in adolescence. There are critical gaps in understanding how and why ACEs are experienced by particular groups of young people and what factors contribute to risk and resilience. This research aims to: a) understand the lived experiences by which ACEs in diverse young people unfold to affect their mental health and b) utilise and understand how novel creative and participatory arts approaches can contribute new knowledge about ACEs to inform future interventions for adolescent mental health.

**Methods and analysis:** An interdisciplinary collaboration, this utilises a blended mixed-methods approach as a triangulation between qualitative research methods, arts-based practice research and a participatory-community-research model. Framework-analysis is utilised as is appropriate to the complexities of data collection in interdisciplinary studies, working across teams, combining analysis of themes across a group (ACE-affected adolescents) with consideration of individual participants through interviews and creative media. This study will recruit 100 young people aged between 10-24 in England in; Cornwall, Kent, Leeds, London and Oxfordshire. This study is part of a larger project by the same authors, investigating adolescent mental health. In this discovery phase, creative practices are being used in conjunction with qualitative methods of data analysis to enable us to elicit, examine lived experience and youth voice as core features of enquiry. This facilitates understanding of the role of creative practices in helping young people share the events in their lives that they feel have been significant in shaping their views of themselves, of others and of their current mental health and wellbeing.

**Ethics and dissemination:** This study is sponsored by the University of Oxford. Ethical approval obtained from institution (R71941/RE001) and NHS Health Research Authority committees. (23/WM/0105) The outputs from this study will be shared, locally, nationally and globally.

**Strengths and Limitations (five):**

- This study aims to understand the lived experiences of adolescents aged between 10-24 regarding adverse childhood experiences (ACEs).
- This study is recruiting adolescents from a range of communities and geographic localities in England, including, rural and urban.
- There is a plethora of research examining the association between ACEs and mental health outcomes, but a paucity of research has employed a blended mixed method approach utilising qualitive research methods, arts-based practice and a participatory community research model.
- This study will purposively endeavour to ensure each adolescent’s voice is heard and the study will engage with those individuals who may otherwise not be able to participate in research.
- This study is focused on recruiting in England and thus the findings it is hoped will be useful to policymakers and clinicians in the devolved nations but the authors recognise that adolescents in the devolved nations may have experienced different ACEs to those adolescents in this study.

## Introduction

Adverse childhood experiences (ACEs) are distressing events that may occur in a young person’s life. These events include, but are not limited to; experiences of psychological, physical or emotional abuse, neglect, loss of parents or siblings. The term ACEs encompasses potentially traumatising events that occur as a result of deliberate intentional harm, as well as events that are non-intentional in nature.

ACEs cluster in socio-economically deprived areas, have mental health impacts across the lifespan, and can even reduce life expectancy by up to 20 years^1^.Three in four ACE-exposed adolescents experience mental health impacts due to the experience of ACEs develop mental health problems by the age of 18, including major depression, conduct disorder, alcohol dependence, self-harm, suicide attempts, and post-traumatic stress disorder ^2^.It is important to note that ACEs do not evolve in a linear pattern method and recent research indicates that adolescence is a point in which the impact of ACEs on mental health becomes evident. Adolescence is a formative stage of development in which environment and individual influences contribute to/ determine mental health outcomes.

The last decade of research shows the devasting impact of early life adversity on an individual’s mental and physical health^3-5^. Childhood maltreatment is a significant risk factor for later psychopathology, as well as neurodevelopmental, social and emotional impacts.^6^The mechanisms through which ACEs lead to these outcomes is less clear, but there seems to be a link between specific pathways and specific symptoms, such as the role of epigenetics, and environmental influences^7^ Other proposed mechanisms include the interaction of pre-natal adversity and postnatal environmental factors^8^. Whilst other researchers have explored stress-related cascades and psychosocial mediating mechanisms, including the impact of attachment theory in the relationship between childhood adversities and severe mental health outcomes^9 10^

The research to date on childhood adversities has predominantly utilised cross-sectional designs and longitudinal designs largely using quantitative methods of collection ^11 12^.There have been few studies that have employed qualitative methods ^13 14^to investigate the role of ACEs on adolescent mental health outcomes but few studies to date have utilised non-verbal/arts based alternatives. The diversity and complexity of people’s experiences of ACEs in addition to the recognised barriers to reporting, require new to capture adolescents’ lived experiences. The research that has been conducted indicates that adolescents who have experienced ACEs are able to recall and reflect on possible adversities^15 16^. There have been far fewer studies that have used creative based approaches to explore adolescents’ lived experiences of possible childhood adversities ^17 18^. This is particularly surprising given the reported impact that creative-based approaches and methodologies has on those who have experienced adversities^19-21^

These facts highlight the importance of research on the experiences of adolescents in today’s society. However, given the limitations of conventional research methods, such as poor response rates to surveys, collective avoidance, and the distress associated with recall of such events, a creative and supportive approach may offer advantages. It is also critical that research exploring adolescents lived experiences is fully informed by youth voice through participatory approaches, UNCRC Article 12 ^22^states children and young people have the human right to have opinions and for these opinions to be heard and taken seriously. This involves working with adolescents as co-producers at every level of the research process from design to delivery of output. Finally, it is essential to ensure that research on ACEs is sensitive to and addresses diverse identities, localities and place contexts and highlight the needs of those who are under-represented in ACE and mental health research. This protocol sets out work package 1 of the ATTUNE project which is a multi-site interdisciplinary project until august 2025.

## Objectives

This multi-site study seeks to understand the lived experiences of adolescents (aged 10-24 years) in England regarding ACEs. Specifically, the study’s primary research questions are:

1. What are the psychological and geo-social-economic contextual mechanisms by which ACEs unfold to affect or safeguard the mental health and lives of YP (aged 10-24)?
2. Do creative and participatory arts approaches facilitate the generation of new and transformative experiential data to improve understanding of mechanisms, and actions for prevention and care interventions?

From these two main research questions we have derived five substantive, and four methodological research questions:

### Substantive Research Questions

1. What are the lived experiences of diverse ACE affected young people?
2. How do young people define what an ACE is and their experience of them?
3. How do those young people affected by ACEs define and explain their mental health?
4. What are the mechanisms through which ACEs unfold to affect an individual’s mental health negatively?
5. What are the prevention and intervention opportunities?

### Methodological questions

1. Do creative and participatory arts approaches help young people to share their experiences?
2. Do participatory arts approachesLengage, empower and support young people, personally and as actors?□
3. Do creative and participatory arts approaches generate new and transformative experiential data to improve understanding and actions for prevention and care interventions? □□
4. What dissemination audiences and strategies are proposed by young people, partners and community agencies?

## Methods and analysis

### Overview of study design

This study is part of the wider ATTUNE project (<u>ATTUNE — Department of Psychiatry (ox.ac.uk)</u> and the first of six workstreams. Diverse groups of young people across the adolescent age range (10-24) will be invited to take part in a series of arts-based workshops across different modalities, led by creative practitioners. During the workshops, participants will engage in creative practice individually or in groups with other young people and guided conversations, to share their lived experiences of ACEs, mental health and current perspectives to whatever degree felt comfortable to them. Workshops will be delivered across five key geographic regions of England; Cornwall, Kent, Leeds, London and Oxfordshire, enabling delivery to take place in rural, coastal and urban environments. As part of their entry into the ATTUNE programme, participants will be invited to complete a survey that characterises the individuals involved with regards to demographic information, psychosocial variables including mental health and experiences of specific ACEs. A post-workshop survey including the psychosocial measures described below, and an open question will be used to understand participants’ experiences of the workshop series and its potential impact.

### Participants

This study will recruit between 80-100 adolescents from across the regions identified. A purposive sampling strategy will be adopted to ensure that the participants recruited are diverse and includes young people across different age groups (10-15, 16-24 years) and gender identities, and from rural, urban, and costal spaces across different regions (study sites). The study will include substantive representations from groups whom are known to experience an excess preponderance of ACEs and to have unique experiences which are less-well documented, i.e. the LGBTQIA+ community, ethnic minorities, refugees and asylum seekers, traveller community, and neurodivergent adolescents. As a research team we are purposively seeking these groups as they are currently not represented within ACE research ^23 24^.To aide recruitment and comply with safeguarding requirements, recruitment of participants will be facilitated through appointed responsible contacts in youth organisations, charities, schools and NHS sites. No general public recruitment will take place.

### Inclusion criteria

Young people must be between the ages of 10 and 24, UK residents, available to commit to the full workshop programme (within the parameters of daily life) and have provided informed consent to participate (or assent plus caregiver consent if under 16 or unable to consent independently). To ensure purposive sampling across identity subgroups we will monitor the characteristics of the sample through a sampling matrix/database.

### Exclusion criteria

The exclusion criteria for this study is that; adolescents younger than 10 and individuals older than 24 years of age, adolescents who do not provide informed consent or assent/caregiver consent to participate.

### Stakeholder involvement in study design

The study is focused on the experiences of young people and their meaningful participation throughout the research. To guide our research process and methods, we will recruit local young people advisory groups (YPAG) and host regular multi-site meetings. The project has appointed an interdisciplinary Advisory Board to include policy makers, clinicians and academics, the NSPCC, National Children’s Bureau and Centre for Mental Health, alongside young people. At the initial stage of the study, we will work with young people to consider ethical aspects of the project, as well as the study design, workshop guide, questionnaires and methods used to elicit sensitive information.

Young people will continue to be involved throughout the life course of the study, including in guiding data analysis, interpretation, dissemination and governance arrangements. YPAG meetings will provide a forum for young people to contribute to aspects of the study and collaborate with the study team. We will aim to ensure appropriate training and support for young people to contribute, and will seek feedback on our involvement strategies. Clinicians will be available from the study team to discuss any issues that may have arisen in a safe environment, as it is acknowledged that this study touches on sensitive issues. Advisory board meetings will be held quarterly in hybrid form to report and receive input into key aspects of the study.

Moreover, to facilitate dissemination, reach and impact, a separate work stream runs in parallel to this workstream that is focused on pathways to impact. This parallel stream includes members of the NSPCC, National Children’s Bureau and Centre for Mental Health to ensure that there is stakeholder involvement at key milestones within the study. This study will therefore liaise with these actors throughout its life cycle.

### Data collection

#### Research setting

Creative arts-based workshops including dance, drama, film, photography and mixed media will be arranged for young people though local organisations in environments that are appropriate for the group due to familiarity (e.g. Schools, community youth groups) or context (e.g. accessible studio spaces on university campuses). Each workshop will be held in person but with options for virtual delivery if necessary (e.g. accommodating communication preferences). COVID resurgence mitigation options are developed and invoked if necessary through the online model. Prior to taking part in the creative workshop, participants (and caretakers where applicable) will be invited to complete a consent form online, or in hard copy where more appropriate, and will have a chance to ask the research team questions. These consultations with potential participants will take place in person.

#### Participant characterisation and workshop evaluation

Once individuals have consented to take part in the ATTUNE Project (across workstreams), they will be asked to complete an entry survey. Questionnaires administered pre-workshops will help characterise those involved in terms of demographic characteristics, certain ACEs and life experiences, including standardised measures of mental health and wellbeing, social connectedness, empowerment, support-seeking intentions, emotion regulation and mental health and wellbeing (see Table 1). Open questions will be used to understand participants’ expectations for the workshop work. The psychosocial questionnaires will be repeated post-workshop to help understand the impact of young people’s participation in the workshops, alongside an open question about perceived impact of participating. These questions are all submitted online and the arts-based researchers do not see the responses.

**Table 1.**
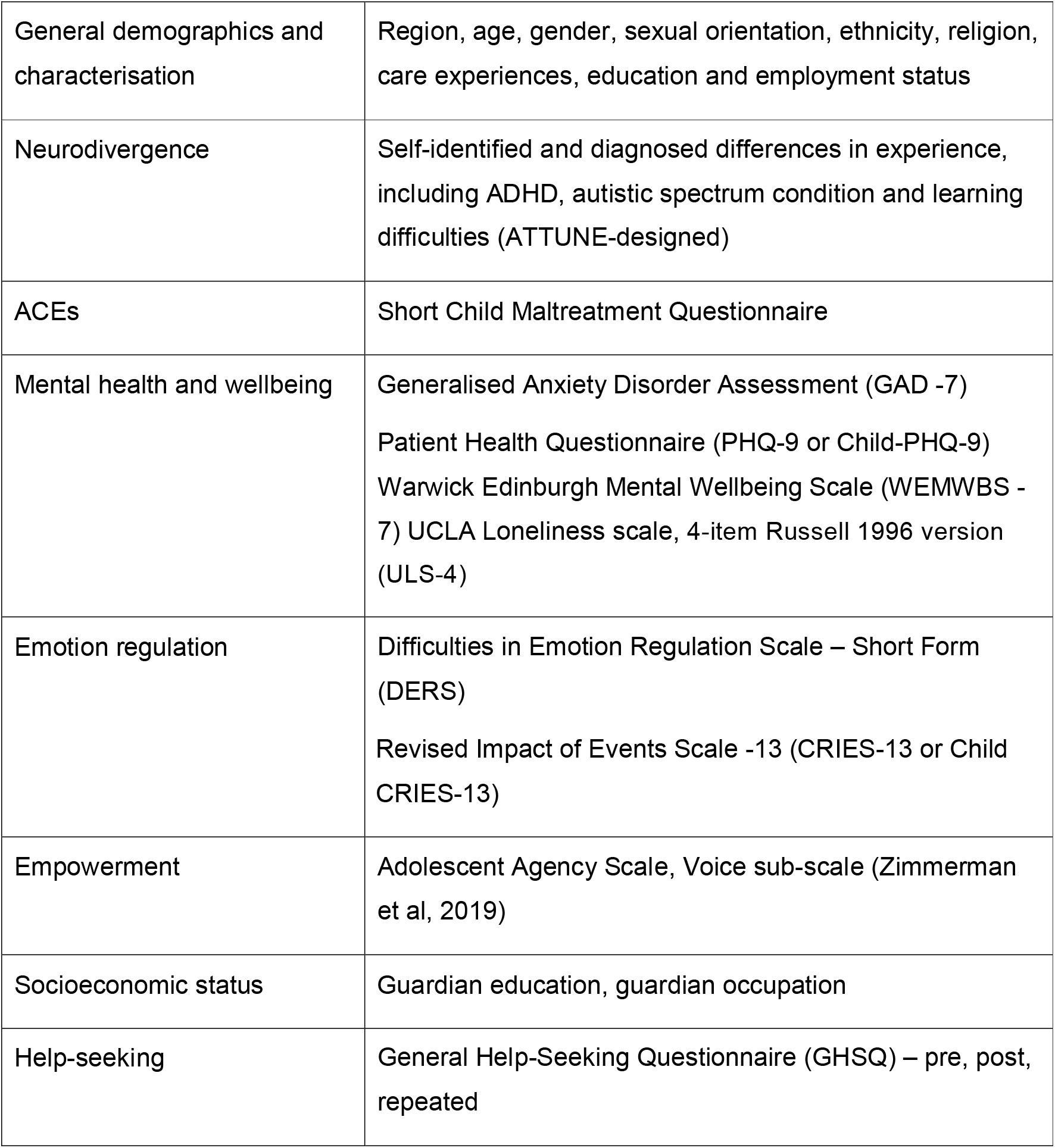
Measures included in the ATTUNE Programme survey.

#### Co-discovery of experience as evidence: data collection using creative methods

The ‘discovery’ data for the ATTUNE project refers to first person experiences and narratives explored through creative practices and resulting mixed media. The design of the methods to explore lived experience is in the context of participatory and socially engaged arts (applied drama, dance, music, filmmaking and amination) and practice research in arts based disciplines^25 26^. Creative practices are often used as an accessible way of engaging marginalised groups in participatory research, offering diverse and flexible modes of communication and representation through visual, auditory, digital, and non-text-based media (drawing, music, movement, photography, film). Working in arts contexts is a means of working holistically in ways that are embodied and sensitive to the complexities of the intersubjective, cultural and environmental realities of adolescent experiences. As the arts are also important to youth culture, they offer appropriate tools for engaging with the lived experience of adolescents and foregrounding youth voice.

A detailed guide for the workshop series is provided in Appendix 1/Supplementary material. The design and methods of this study have been carefully planned with specialist mental health practitioners, young people and a research team that includes people with lived experience of mental health challenges, adverse childhood experiences, and those who identify as LGBTQ+, neurodivergent, or from diverse ethnic groups. Both content and delivery are informed by the previous work of team members using arts based approaches and participatory research to investigate adolescent mental health; photovoice^27^, participatory film ^28^interactive digital media^29^, and socially engaged drama, theatre and music as multimodal practices^26 30-32^including blended arts and psychology programmes.

In the current study, a variety of arts practices are used, including performance (e.g., drama, theatre and dance), still and moving visual art (e.g., drawing, zine design, photography, film and animation) and writing, as part of a multimodal approach. Multimodal creative methods generate rich verbal, visual and bodily data, appropriate to the richness and complexity of ACE-affected adolescent experience and the diversity of the participating groups. This is a novel approach as most arts-based health interventions are grounded in a singular arts discipline thereby building an evidence base for the efficacy of a particular creative practice with a particular client group. In the ATTUNE project, however, arts modalities were specific to the contexts in which data was being collected. These vary from participatory film, photography, film animation and improvised drama, to dance based practice involving ‘relational choreography’ to produce a film blending movement, spoken word and documentation of participants’ practical process Other groups engage with an explicitly multimodal programme in which themed activities (safe creative space, masks and shields) are explored through mixed media (creative writing, collage, action art). Collectively, the arts practices combine image based, text based (verbal and narrative) and embodied (performance based) modes of data assemblage^33^

Whilst the art practices accommodate diversity of participants and form, continuity is created through the structures of the practical workshops. These are designed in relation to the research questions, using a thematic approach with prompts (“creative conversations”) as a scaffolding for the activities. For example, in the first session of the multi-model workshops, the focus on safe creative space generates discussion of how safety is experienced by the participating young people and what they consider to be risks and threats, as well as considerations of how place impacts on adolescent mental health. Subsequent sessions involve prompts related to senses and memory, objects and emotion, and storytelling and journeys with opportunities for working individually and in groups to engage with research questions through the structure of creative conversations. The data assembled through this process is a triangulation of transcripts (recorded conversations responding to prompts), art work and field notes. The creative conversations have a similar function to semi structured interviews, but the creative context offers a space in which young people are more open about their experiences ^34-36^.This is due to the creative environment building trust as well as its liminal qualities as a space associated with freedom, multi-sensory self-expression and play. This is also key to the therapeutic benefits of such practices^37^.

Workshops are conducted over six to eight weeks dependant on the cohort, relationship building and timing of delivery, working with and through the key organisations involved in the project (e.g., schools and school timetables). In some sites, workshops may run as intensive, studio-based programmes. It is crucial that the workshops run at the pace of the participants, as forming relationships with the practitioner is crucial to the success of creative based workshops. The series of recorded key conversations that accompany each workshop invite participants to express, explore and reflect on themes related to mental health and life experiences, in a creative arts-based, non-judgemental context.

The multimodal method results in an *assemblage* of data (recordings, transcripts, fieldnotes, art work), requiring an interdisciplinary approach appropriate to this complexity, to ensure rigour whilst respecting richness.

### Data analysis

#### Qualitative data

Our analysis will focus on addressing the substantive and methodological research questions we have set out for this project.

The decision was taken to analyse the dataset thematically using an adaptation of the Framework staged approach ^38^.This has a series of stages (i) familiarisation, (ii) coding (identifying categories and themes to create a codebook); (iii) Indexing (developing and applying the thematic framework to the transcripts); (iv) charting and summarising using a matrix; (v) interpretation and mapping; (vi) indexing with narratives and art materials for (1) findings specific to or generalisable across groups and settings, (2) and resiliency mechanisms, prevention and intervention opportunities.

For the ATTUNE study, this approach offered a series of benefits: firstly, it is a step by step technique, primarily used in applied research, that is appropriate for multi-disciplinary teams and can be taught to novice researchers or community participants engaged in the analysis of lived experience accounts. It combines deductive and inductive approaches which means it can be used pragmatically so that, for example, the categories (deductive) can be aligned with the research questions whilst also enabling an inductive and intuitive approach through the thematic codes arising from close analysis of data. Secondly, the matrix facilitates a focus on individuals as the charting and summarizing means the researcher considers how themes occur across transcripts (columns) whilst still being able to consider particular participants (row). Thirdly, the method is dynamic, so popular for its versatility and flexibility as researchers can make changes throughout the process in ways that complement the iterative nature of practice research. Finally, it offers a systematic approach that is appropriate for a complex project design.

A series of strategies are in place to ensure the trustworthiness of the findings in the qualitative analysis:

1. There is cross disciplinary representation in the team contributing to the analysis. Each set of transcripts is being coded by at least one researcher who was engaged in delivery of the workshops, and this is counterbalanced by the perspectives of at least one researcher who was outside the process. The inclusion of practitioner and participant perspectives in the analysis is a distinctive feature of the approach, while the balance of insider and outsider perspectives mitigates the risks of personal biases influencing the findings.
2. A detailed diary of the process and decisions is kept online to ensure there is a record trail.
3. Cross site comparisons to identity similarities and differences between accounts and to ensure different perspectives are represented.
4. The inclusion of different forms of data as an assemblage of forms and perspectives (e.g., practitioner field notes and recordings of workshop debriefs). This ensures the analysis is supported by the richness of thick description and practitioner observations ^39^
5. Respondent validation involves us inviting participants to comment on our findings and whether the themes and concepts adequately reflect their experiences.
6. A process of data triangulation brings the different materials and perspectives into dialogue and will engage participants, researchers and the public audiences for the work. In order to combine the narrative and art data to inform learning and to ensure the artwork is valued for its affective qualities, a process of creative evaluation will be conducted as part of the triangulation process. The data assembled will include perspectives from the arts practitioners on the knowledge emerging from the artwork produced, explanations and interpretations from young people as makers and audiences for the artwork as well as critical analysis of the artefacts, informed by the framework categories and coding. An evaluation tool kit will be co-produced with the young people to ensure that youth voice is at the heart of this process.

#### Survey data

Descriptive statistics will be used to characterise the sample with regards to age, gender, sexual orientation, ethnicity, socioeconomic status, place of residence (region and rurality), ACEs experiences and frequency, care experiences, employment and education status, mental health and wellbeing, help-seeking, empowerment and emotion regulation. This will be used to contextualise the qualitative analysis. Post-workshop survey data, including the open question on impact, will complement young people’s reflections of their experience of the arts-based workshops, collected throughout and particularly at the end of the workshop series.

## Ethical considerations

The workshop programmes have been carefully designed, considered ‘trauma-informed guidelines for practice, to ensure safe and comfortable environments for young people given the sensitivity of the workshop content (e.g., check-ins, trust-building activities and follow up between workshops). Adjustments to the programme will be made to facilitate inclusion and participation from different groups (e.g., those with learning disabilities, non-English speakers)

All arts-based practitioner and researchers in contact with participants are DBS checked, have undergone safeguarding training and have significant experience working with and supporting vulnerable groups. Each of the sites will have a safeguarding lead to ensure appropriate and time-sensitive responses to any identified risks and according to an approved safeguarding protocol. We will be providing practitioners with a ‘creative reflective practice’ space run by an independent clinician to support practitioner wellbeing and safe practice (delivered by author LS)

A pioneering feature of this study’s methodology is the dissemination of selected artwork and through a range of communication methods and platforms, honouring young participants’ right to be recognised as the authors of their art work. The process of curating participants’ artwork will be conducted sensitively and written and recurrent verbal consent will be sought for dissemination. To acknowledge their contributions as artmakers, participants will be invited to both be named as the author of their artworks, to use a pseudonym or to remain anonymous. As part of this consenting process participants will also be encouraged to consider the right to be forgotten and their potential future viewpoints

The team has published ethics guidance on arts-based methods for young people who experienced ACEs (see ^35^) Throughout the project we aim to continue generating new ethics knowledge and guidance to support future participatory arts projects.

## Dissemination

A particular novel output of this study is the creation of an art portfolio that captures the perspectives and stories of participants with the arts-based data insight triangulated by young people. A triangulation approach will be used which will involve referring to the codebook and employing This will include videos of performances, animation, artefacts, films and zines as well as digital versions of the multimodal outputs and documentation of the creative process. The artwork will be displayed throughout the life cycle of the study (2022-2025) through curated exhibitions, screenings and publications. This will involve exhibitions in the regions where workshops have taken place as well as across the four nations of the United Kingdom.

The research team will disseminate emergent insight throughout the study on social media platforms including but not limited to Twitter, LinkedIn, Instagram and TikTok. These platforms will be monitored by two young people who are part of the study team and thus will ensure young people’s views remain at the very centre of the study.

Research papers, posters and seminars talks will be given at relevant conferences both in the public and private sectors to ensure that emergent insight is disseminated through appropriate forums to policymakers, clinicians and members of the public. Results will also be shared in peer-reviewed journal publications, co-authored by members from other work streams and young people where appropriate. Findings from this work-package will inform the design of digital resources such as a documentary and a game.

Furthermore, hybrid knowledge exchange conferences will be organised for individuals from a range of professions and sectors, including but not limited to, mental health professionals, psychotherapists, social workers, education professionals, commissioners and youth workers. These will occur at intervals throughout the lifetime of the project. The aim of these conferences will be to share findings from the study on experiences, coping and help-seeking and to showcase how creative arts modalities can help empower, engage, understand and intervene to improve youth mental health in specific contexts for specific identity groups.

## Data Availability

No data is available.

## Funding

This work was supported by the UKRI Medical Research Council. (MR/ W002183/1).

## Patient and public involvement

Patients and/or the public were involved in the design, or conduct, or reporting, or dissemination plans of this research. Refer to the Methods section for further details.

## Patient consent for publication

Not applicable

## Competing interests

None declared.

## Acknowledgements

The authors wish to thank each member of the wider ATTUNE research team who has helped in the design, conceptualisation of the project including those delivering those delivering the workshops in each location and the broader team.

## Contributors

KB is the principal investigator for the ATTUNE study(<u>ATTUNE — Department of Psychiatry (ox.ac.uk)</u>). KB, IB, AMW, SHJ, PC, BT, NS, and GP all made substantial contributions to the conceptualisation and design of the study. AMW, NS, KB, GP, BT, PC, LS, SHJ and IB drafted the first version of the manuscript. All named authors reviewed the manuscript for intellectual content, approved the final version to be published and agreed to be accountable for all aspects of the work.

## Notes

### Competing Interest Statement

The authors have declared no competing interest.

### Funding Statement

This study is funded by the UKRI-MRC, grant reference(MR/ W002183/1).

### Author Declarations

This study is sponsored by the University of Oxford. Ethical approval obtained from institution (R71941/RE001) and NHS Health Research Authority committees. (23/WM/0105)

